# Predicting intentions towards long-term antidepressant use in the management of people with depression in primary care: A longitudinal survey study

**DOI:** 10.1101/2024.02.16.24302927

**Authors:** Rachel Dewar-Haggart, Ingrid Muller, Felicity Bishop, Adam W A Geraghty, Beth Stuart, Tony Kendrick

## Abstract

**Background:** Over the last two decades, antidepressant prescribing in the UK has increased considerably, due to an increased number of people staying on antidepressants for longer. Even when treatment is no longer clinically indicated, qualitative research suggests many people continue due to a fear of depressive relapse or antidepressant withdrawal symptoms. The quantitative effects of peoples’ beliefs and attitudes towards long-term antidepressant use remain relatively unexplored.

**Objectives:** To determine the extent to which beliefs and attitudes towards antidepressant treatment predict intentions to stop or continue long-term use; and whether intentions translate into actual discontinuation.

**Methods:** A questionnaire survey formed the main component of an embedded mixed-methods study. Twenty general practices posted questionnaires to adults aged over 18 receiving continuous antidepressant prescriptions for over two years. Outcomes and predictors were determined using an extended model of the Theory of Planned Behaviour, conducting exploratory descriptive and regression analyses. The primary outcome was participants’ intentions to discontinue antidepressants. The secondary outcome of behaviour change was determined by any change in antidepressant dosage at six months.

**Results:** 277 people were surveyed from 20 practices, with 10 years median antidepressant duration. Mean questionnaire scores for intention and subjective norms towards starting to come off antidepressants were low, and 85% of participants declared that continuing their antidepressant was necessary. Prescribing outcomes retrieved from 175 participants’ medical records six months after they completed the survey found 86% had not changed their antidepressant, 9% reduced the dose, only 1% discontinued their antidepressant, and 4% increased the dose. More favourable attitudes towards stopping, and normative beliefs about depression, were the strongest predictors of intentions to stop long-term antidepressant treatment.

**Conclusion:** Given few intentions to stop taking antidepressants, patients should be made more aware of the importance of ongoing antidepressant monitoring and review from their primary care practitioners. This would promote discussion to support an attitudinal change and initiation of antidepressant tapering where appropriate.

## Introduction

Over the past two decades, antidepressant prescribing rates have risen considerably, nearly doubling between 2008 and 2018.[1, 2] Between 2015 and 2018, the rate of antidepressant prescribing in primary care increased from 15.8% to 16.6%[3, 4]; with 7.3 million people prescribed antidepressants in 2017/18, at an annual cost of approximately £266 million.[5] The considerable rise in the volume of antidepressant prescribing in primary care is due to an increased number of people receiving continuous antidepressant treatment for longer.[2, 6-11]

A third to a half of people taking long-term antidepressants may have no evidence-based indications to continue treatment, and could try to stop.[12] Long-term outcomes of antidepressant-treated depression are generally poor,[13] and antidepressants may additionally pose the risk of adverse long-term iatrogenic effects such as sexual problems, weight gain, feeling emotionally numb and the perception of being addicted to medication.[14–17] In people over 65, adverse effects associated with antidepressant use include falls, seizures, strokes, low blood sodium, and cardiac arrhythmias.[18]

People with a stronger belief in the effectiveness of medication are more likely to be taking antidepressants, more likely to believe that their condition has a chronic timeline, and more likely to be currently depressed.[19–21] Individuals have a greater perceived need for antidepressants if they believe their depression is caused by chemical imbalances or is hereditary.[22, 23] These findings suggest that a greater belief in the chronic and biochemical nature of depression will lead to longer-term antidepressant treatment, as patients may believe that pharmacological interventions are more effective at symptom management than non-drug treatments. However, higher self-efficacy in managing depressive symptoms and a belief in using talking therapies or engaging in activities such as exercise or keeping busy to manage depression is associated with improved depression outcomes.[19, 24]

While there is considerable qualitative evidence on the issues surrounding ongoing antidepressant treatment in primary care, the quantitative effects of peoples’ beliefs and attitudes towards long-term antidepressant use remain relatively unexplored.

The aims of this study were to investigate the extent to which beliefs and attitudes towards depression and antidepressant treatment predict intentions to stop or continue long-term antidepressant use; and whether these intentions translate into actual behaviour of antidepressant discontinuation. A further objective was to determine how well participants’ beliefs and attitudes can be predicted by the Theory of Planned Behaviour (TPB).[25]

## Materials and methods

### Study Design

A cross-sectional, mixed-methods design using a quantitative questionnaire survey study with nested qualitative interview study was used, with data collection occurring concomitantly.[26, 27] The findings from the qualitative study will be reported elsewhere. A copy of the study protocol is provided as Appendix S1.

### Setting

Twenty group general practices from the Clinical Research Network (CRN): Wessex and CRN: West of England were recruited to the study from November 2017. Participant recruitment began in February 2018 and ended in February 2019.

### Participants

Practices were asked to conduct a database search to identify patients over the age of 18 who had been continuously receiving antidepressant prescriptions for two years or longer. Practices were given both a list of British National Formulary (BNF)[28] antidepressant names and Read codes[29] for diagnoses and symptoms of depression to conduct the search. Patients were excluded if they were prescribed antidepressants for conditions other than depression, had a comorbid psychiatric condition or depression managed in secondary care, or were terminally ill, lacking capacity, or deemed unable to take part after screening by a GP. Participants were not excluded based on their severity of depression or if they had any comorbid physical conditions.

Eligible participants were sent an invitation pack in the post by their GP practice. Each practice was asked to send packs to up to 140 patients. The pack included an information sheet, questionnaire booklet, and consent form. The study was ethically approved through proportionate review by Yorkshire & The Humber – Leeds East Research Ethics Committee (REC ID: 17/YH/0223). Completion of the questionnaire indicated implied consent.[30] However, participants were required to provide written consent for their GP practice to provide anonymised medical notes data. The authors did not have direct access to participants’ medical records, and only data that were relevant for the outcome of the study (antidepressant prescribing data and/or any record of consulting with a primary care practitioner for a mental health review within six months of the participant completing the questionnaire) were obtained. Requests for this data from the GP practices were made up to March 2020, due to the Covid-19 pandemic affecting practices’ capacity to continue research.

### Outcomes and variables

Outcome and predictor variables were determined by using an extended model of the (TPB) (**Error! Reference source not found.**).[25] The primary outcome was participants’ declared intentions to start to come off antidepressants. The secondary outcome of actual behaviour change was determined by a reduction in antidepressant prescription dosage or attending an appointment to discuss possible discontinuation within the six-months of completing the questionnaire. Further constructs of salient beliefs,[21, 31-35] past behaviour,[31, 36] symptom severity,[37] and current antidepressant duration[21, 33] were hypothesised to predict intentions to stop or continue long-term antidepressant use and added to the model.[25, 31, 38-40]

**Fig 1.**
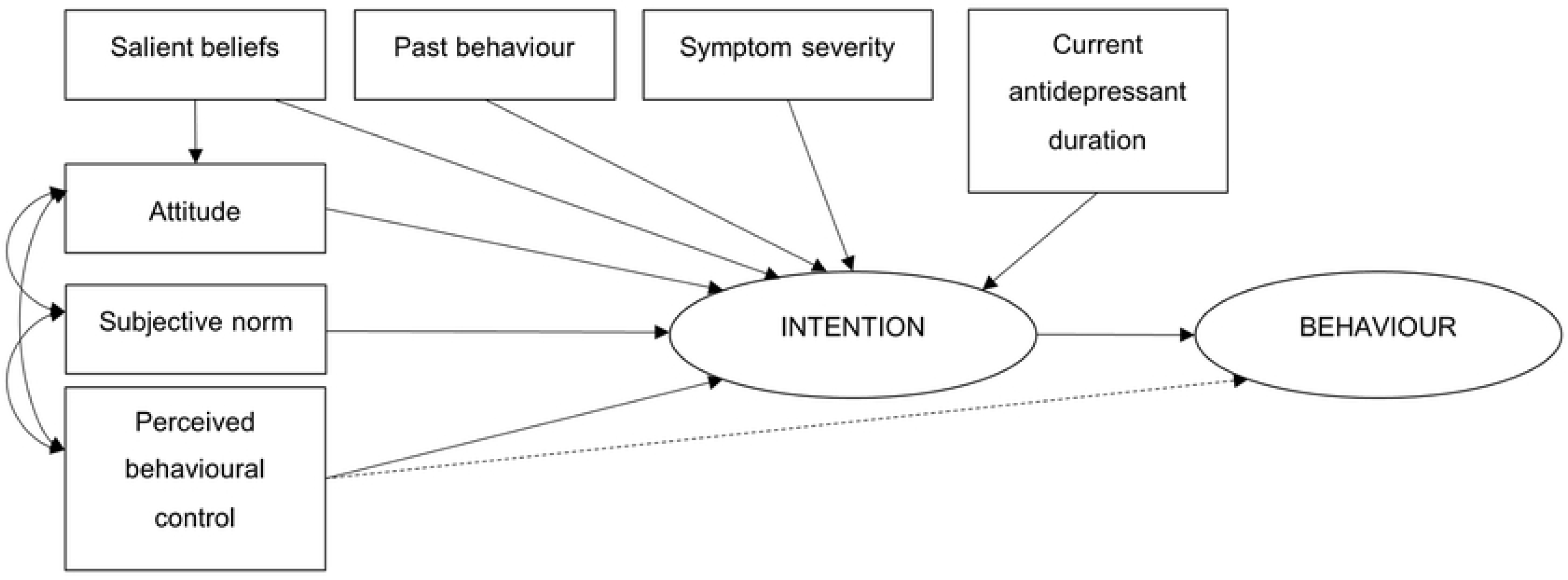
An extended model of the Theory of Planned Behaviour.

### Data measurement

The questionnaire survey participants were asked to complete is included as Appendix S2. The medical notes data form that practices were asked to complete is included as Appendix S3.

#### Demographic characteristics

Participants provided demographic characteristics including gender, age, ethnicity, marital status, number of dependants, level of education, and occupation.

#### Intention, attitudes, subjective norms, and perceived behavioural control

Intention, attitudes, subjective norms and perceived behavioural control (PBC) towards starting to come off antidepressant treatment were measured using Likert rating-scale items, created using guidance for developing questionnaire items based on the TPB.[25, 41] Items were developed and refined through cognitive interviews as part of RDH’s PhD.[42] Items with Cronbach’s alpha correlations of α >.60 suggested good internal consistency.[41, 43] Composite variables for the direct measures were calculated by creating a mean score for the items relating to intention, attitude, subjective norm, and PBC, with higher scores indicating stronger beliefs. Additional descriptive statistics around attitudes towards antidepressant discontinuation were measured using items from the Patient Attitudes towards Deprescribing (PATD) Questionnaire.[44]

#### Salient beliefs

Necessity and concern beliefs about antidepressants were measured using the Beliefs about Medicines-Specific Questionnaire (BMQ-Specific).[45] Items were modified by changing the word ‘*medicines*’ to *‘antidepressants’*.[35] A total score of both necessity and concern items were calculated and interpreted as continuous scales, with higher scores indicating stronger beliefs in the necessity of, or greater concerns about taking antidepressants. *Physical cause*, *chronic timeline,* and *medication to control/cure* variables of the Beliefs about Depression Questionnaire (BDQ)[45] were calculated by mean scores from 6-point Likert rating scales, where higher scores indicated stronger beliefs.

#### Past behaviour, current treatment duration and symptom severity

Participants provided dichotomous data for past behaviour items adapted from the PATD Questionnaire.[44] Participants provided self-report data on the current duration of their antidepressant treatment. The Patient Health Questionnaire (PHQ-8)[46] was included to measure current symptom severity of depression. The PHQ-8 includes the same items as the PHQ-9,[47] but excludes question nine, which assesses thoughts of harm or suicidal ideas.

The omission of this item only has a small effect on scoring, and identical thresholds are used for both the PHQ-8 and PHQ-9 questionnaires.[46, 48] Symptom scores are categorised to five levels of severity: minimal (0-4), mild (5-9), moderate (10-14), moderately severe (15-19), and severe (20-24).

#### Behaviour

Medical record reviews were carried out to measure the proportion of participants who consulted with a health professional at their GP surgery to review their mental health and determine whether they had started to discontinue treatment, indicated by a reduction in their prescribed antidepressant dosage. Participants were categorised into two groups based on their prescribing data: reduced (reduced or stopped) and did not reduce (increased, no change, or changed antidepressant type).

A summary of all predictor variables included in the model, along with the questionnaire items used to measure each predictor variable, are described in Table 1.

**Table 1.** Predictor variables included in the extended model of the Theory of Planned Behaviour.

| Variable | Description | Survey Measure |
| --- | --- | --- |
| Intention | Intentions towards starting to come off antidepressants in the next six months | TPB |
| Attitude | Attitudes around the benefits and disadvantages of starting to come off antidepressants in the next six months | TPB |
| Subjective norm | Perceived opinions of others about starting to come off antidepressants in the next six months | TPB |
| PBC | Perceived self-efficacy and controllability around starting to come off antidepressants in the next six months | TPB |
| Salient Beliefs |  |  |
| *Necessity* | Beliefs around the necessity of antidepressants | BMQ-Specific |
| *Concerns* | Concerns about taking antidepressants | BMQ-Specific |
| *Medication* | Beliefs around the need for antidepressants to control/cure depression | BDQ |
| *Physical* | Beliefs around the physical causes of depression (genetics, illness, chemical imbalance) | BDQ |
| *Chronic* | Beliefs around the chronic timeline of depression | BDQ |
| Past Behaviour |  |  |
| *With doctor* | Stopping previous antidepressant treatment with a doctor's knowledge | PATD Questionnaire |
| *Without doctor* | Stopping previous antidepressant treatment without a doctor's knowledge | PATD Questionnaire |
| *Successfully stopped* | Duration of previous symptom free episode while off antidepressant treatment | Bespoke item |
| Current antidepressant duration | Current duration of antidepressant treatment | Bespoke item |
| Symptom severity | Current depression symptom severity | PHQ-8 |

#### Study size

Guidance on questionnaires using constructs from the TPB suggest a sample size of 80 participants, if a moderate effect size of 0.3 is expected following multiple regression analysis.[41, 49] A ‘rule of thumb’ sample size estimate was calculated based on Green’s procedure; accounting for the potential of a small effect size and potential of overfitting.[50, 51] Approximately 405 participants would be required for a multiple regression analysis and was deemed feasible to obtain in a primary care setting, assuming a 10% response rate.

### Statistical methods

Data were analysed using SPSS version 26.[52] Frequency distributions and means were calculated for participant characteristics and information on participants’ antidepressant use and history of depression. A Pearson correlation was conducted to determine whether there was an association between beliefs around the necessity of and concerns around antidepressant treatment.

Multiple linear regression with robust standard errors was conducted to determine whether Salient beliefs predicted attitudes towards antidepressant discontinuation. Hierarchical regression analysis was conducted to determine whether the addition of past behaviour, current symptom severity, and current antidepressant use predicted intentions to stop antidepressants over and above constructs from the TPB. A binomial logistic regression was anticipated to determine whether intentions and PBC towards starting to come off antidepressants predicted behaviour (indicated by a reduction in antidepressant dose) and to ascertain the effect of intentions to start to come off antidepressants on whether participants had at least one appointment with a health professional. However, nine of the 12 participants who reduced their antidepressants had studentized residuals ±2.5, which were not corrected when conducting a transformation of the variables. Examining the data suggested they were outliers as they all had low intention scores yet reduced their antidepressants six months after completing the questionnaire. Data analysis was conducted using complete cases.

## Results

### Participants

Recruitment of participants is shown in Fig 2. Most patients approached were female (n= 1288, 70.9%), with a mean age of 55.5 years (SD= 15.3, range= 20-96). Three hundred and ninety-seven responses were received (16.9% of those approached). Questionnaires from 120 respondents were excluded from the study, with 68 excluded based on the self-report item for current antidepressant duration. Forty respondents reported antidepressant treatment duration of less than two years, 13 did not provide any information, and 15 provided data that were unclear, for example: “*don’t know*”, “*can’t remember”*’, or “*years”*. One person returned the questionnaire but later requested to withdraw from the study, including their questionnaire data, with no reason given. Two hundred and seventy-seven participants (11.8% of those approached) were entered into the study, and medical data of 189 participants was received (8.0% of those approached). One participant who completed the questionnaire online entered their Participant ID number incorrectly and could not be linked to a practice to request their notes review data. Due to the Covid-19 pandemic affecting practices’ capacity to continue research, requests for patient data ceased in March 2020. Four practices did not return notes reviews.

**Fig 2.**
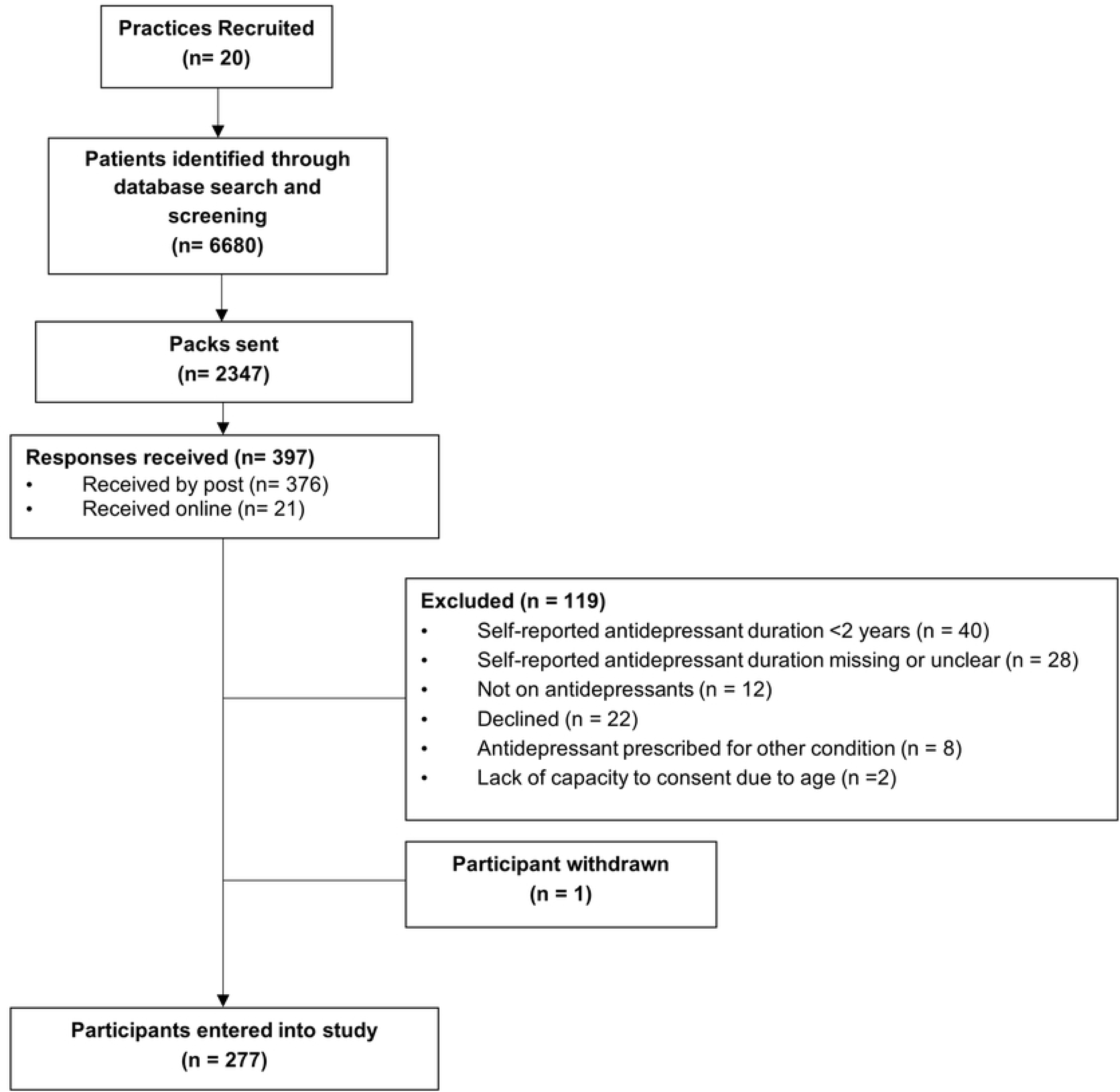
Flow diagram showing participant recruitment.

### Descriptive data

Participant characteristics are provided in Table 2. One participant did not complete the demographic questionnaire. Most respondents were female (n=187, 67.5%) and the mean age was 57.2 years (SD= 14.6). The sample was predominantly white (n= 273, 98.5%), married or cohabiting (n= 190, 68.6%), and in employment (n= 142, 51.3%). There was no significant difference between the mean age of respondents and non-respondents (55.4 years), *t*= 1.43, *p*= 0.15, 95% CI −0.53, 3.38; or percentage of female respondents (67.5%) and non-respondents (72.0%), *χ*^2^= 2.35, *p*= 0.13.

**Table 2.** Participant characteristics.

| Variable |  | N (%) |
| --- | --- | --- |
| Age | Mean years (SD) | 57.2 (14.6) |
|  | Range | 23.3 – 92.5 |
| Gender | Female | 187 (67.5%) |
|  | Male | 90 (32.5%) |
| Ethnicity | White | 273 (98.6%) |
|  | Black Caribbean | 1 (0.4%) |
|  | Asian British | 1 (0.4%) |
|  | Mixed Race | 1 (0.4%) |
|  | Missing | 1 (0.4%) |
| Marital status | Married/Cohabiting | 190 (68.6%) |
|  | Separated/Divorced | 33 (11.9%) |
|  | Widowed | 19 (6.9%) |
|  | Single | 34 (12.3%) |
|  | Missing | 1 (0.4%) |
|  | Households with dependents at home | 120 (43.3%) |
| Education level | None | 30 (10.8%) |
|  | CSE/NVQ Level 1 | 16 (5.8%) |
|  | GCSE/O Level/NVQ Level 2 | 47 (17.0%) |
|  | A Level/BTEC/NVQ Level 3 | 38 (13.7%) |
|  | HNC/HND/City & Guilds/Teaching Qualification/NVQ Level 4 | 36 (13.0%) |
|  | Degree/Higher Degree/NVQ Level 5 | 62 (22.4%) |
|  | Vocational Qualification | 37 (13.4%) |
|  | Missing | 11 (4.0%) |
| Work status | Employed (Full/Part time/Self-employed) | 142 (51.3%) |
|  | Volunteer | 5 (1.8%) |
|  | Unemployed | 6 (2.7%) |
|  | Permanently Sick/Disabled | 21 (7.6%) |
|  | Retired | 84 (30.3%) |
|  | Homemaker | 15 (5.4%) |
|  | Student | 2 (2.6%) |
|  | Missing | 2 (2.6%) |

Characteristics around antidepressant use and depression are presented in Table 3. The most prescribed antidepressants were Citalopram (n= 88, 31.7%) and Sertraline (n= 74, 26.7%). Participants self-reported a median current antidepressant duration of 10 years (IQR 12).

**Table 3.** Antidepressant treatment history and depression symptom severity.

| Variable |  | N (%) |
| --- | --- | --- |
| Current antidepressant | Amitriptyline | 5 (1.8%) |
|  | Citalopram | 88 (31.7%) |
|  | Clomipramine | 2 (0.7%) |
|  | Dosulepin | 1 (0.4%) |
|  | Duloxetine | 6 (2.2%) |
|  | Escitalopram | 5 (1.8%) |
|  | Fluoxetine | 33 (11.9%) |
|  | Lofepramine | 4 (1.4%) |
|  | Mirtazapine | 18 (6.5%) |
|  | Paroxetine | 11 (4.0%) |
|  | Sertraline | 74 (26.7%) |
|  | Trazadone | 1 (0.4%) |
|  | Venlafaxine | 25 (9.0%) |
|  | Missing | 4 (1.4%) |
| Age first prescribed antidepressants (N= 262) | Mean, Median (SD) | 39.4, 38.00 (15.42) |
| Current antidepressant duration (months, N= 277) | Mean, Median (SD) | 134.5, 120.0 (114.52) |
| Previous attempts to stop (N= 277) | With doctor's knowledge | 127 (45.8%) |
|  | Without doctor's knowledge | 97 (35.0%) |
| Successful discontinuation (N= 270) |  | 81 (30%) |
| Duration antidepressant-free (months, N= 71) | Median (SD) | 36.00 (123.16) |
| Current depression symptom severity (N= 255) | Mean, Median (SD) | 8.9, 8.0 (6.81) |
|  | Minimal (0-4) | 83 (30.0%) |
|  | Mild (5-9) | 63 (22.7%) |
|  | Moderate (10-14) | 49 (17.7%) |
|  | Moderately severe (15-19) | 34 (13.3%) |
|  | Severe (20-27) | 26 (10.2%) |
|  | Missing | 22 (7.9%) |

The mean score for symptom severity was 8.90 (95% CI 8.06, 9.74). A higher proportion of participants reported a score of 9 or lower (n= 146, 57.3%), indicating mild to minimal depression symptom severity. More participants had attempted to stop taking antidepressants with their doctor’s knowledge (n= 127, 45.8%). Ninety-two participants (33.2%) had not attempted to stop taking antidepressants at all, compared to 39 participants (14.0%) who had tried to come off antidepressant both with and without their doctor’s knowledge. Eighty-one participants (30%) reported successfully stopping antidepressants in the past, with 71 (87.7%) reporting a median treatment-free duration of 3 years (IQR 8.8) before restarting treatment.

### Outcome data

Prescribing outcomes were recorded at six months for 175 participants (63.2%) (Table 4). The majority (n= 153, 87.4.%) did not change their antidepressants dose, compared to 16 participants (9.1%) who reduced their dose or stopped altogether.

**Table 4.** Prescribing data at six months taken from medical notes reviews.

| Outcome | N (%) |
| --- | --- |
| Change in prescription (N= 175) |  |
| Increase | 4 (2.3%) |
| No change | 153 (87.4%) |
| Reduce | 14 (8.0%) |
| Stopped | 2 (1.1%) |
| Changed antidepressant type | 2 (1.1%) |
| Prescription request method (N= 179) |  |
| Appointment | 26 (14.5%) |
| Reception | 106 (59.2%) |
| Online | 42 (23.5%) |
| Telephone | 3 (1.7%) |
| Repeat box | 2 (1.12%) |

Of the 14 participants who reduced their dose, 11 had a face-to-face appointment with their GP, and one participant had a medication review with a pharmacist. One participant did not have any appointments with a health professional, and no data were provided for the final participant. For the two participants that stopped completely, one had a face-to-face appointment with their GP, and the other stopped requesting antidepressant prescriptions. Fifty-two participants (29.7%) had a face-to-face appointment and eight (4.6%) had a telephone appointment with their GP. Two participants who did not change their antidepressant dose had a medication review with a pharmacist.

### Beliefs about depression and antidepressant discontinuation

Mean scores for beliefs and attitudes towards depression and antidepressant discontinuation are shown in S1_Table. The mean scores for intention (*M*= 2.44, 95% CI 2.23, 2.65) and subjective norms (*M*= 2.35, 95% CI 2.21, 2.49) towards starting to come off antidepressants were low. Pearson’s correlation showed a weak but significant negative correlation between the necessity and concerns of antidepressant treatment (*r*= −0.15, *p*<0.05). Higher mean scores indicated stronger beliefs that depression was *chronic* (*M*= 4.65, 95% CI 4.46, 4.84) and *medication* was needed to help control/cure depression (*M*= 5.12, 95% CI 4.96, 5.28).

Most participants were comfortable with taking antidepressants (n= 134, 84.8%), and nearly all participants (n= 248, 90.2%) agreed that they understood why they were prescribed antidepressants. Conversely, most participants disagreed that they were taking antidepressants they no longer needed (n= 189, 68.5%) or that their antidepressants were giving them side effects (n= 163, 59.0%). Participants showed uncertainty around whether they would like to stop taking their antidepressants (n= 67, 24.2%), or their willingness to stop taking antidepressants if their doctor said it was possible (n= 100, 36.2%). However, if participants were to start to come off antidepressants, over half (n= 167, 60.3%) reported they would be comfortable if their doctor were involved with the process as well as providing follow-up compared to a nurse practitioner (n= 109, 39.4%) or pharmacist (n= 161, 58.1%). Most participants (n= 240, 87.3%) indicated a preference for face-to-face follow-up appointments with their GP.

### Salient beliefs in predicting attitudes towards discontinuation

Multiple linear regression was conducted for 173 participants to determine whether Salient beliefs predicted attitudes towards antidepressant discontinuation, along with the mean and standard deviations (S2_Table). The multiple correlation coefficient (*R*= 0.71) showed a moderate to strong linear relationship between salient beliefs and attitudes towards stopping antidepressants. The proportion of variance in attitudes accounted for by the regression model was *R*^2^= 49.7% with an adjusted *R*^2^ of 48.2%, suggesting a medium effect size.[49]

The coefficients for each of the predictor variables are shown in S3_Table. The slope coefficients show that stronger beliefs in the necessity of antidepressants, along with stronger beliefs that depression can be cured/controlled by medication, has a physical cause, and chronic timeline were significantly associated with more negative attitudes towards stopping antidepressants. Necessity of antidepressants had the largest contribution in predicting attitudes towards discontinuation *t*(167)= −6.80, *p*< 0.001. However, concerns about antidepressants did not significantly predict attitudes towards stopping antidepressant treatment (*B*= 0.04, 95% CI −0.01, 0.09, *p*= 0.06). Overall, the model showed that salient beliefs significantly predicted attitudes towards discontinuing antidepressants, *F*(5, 167)= 33.03, *p*<0.001, adj. *R*^2^= 0.48.

### Predicting intentions

The hierarchical regression analysis for predicting intentions was run on complete data from 161 participants. The means, standard deviations and correlations between variables are shown in S4_Table. Most variables had a significant linear relationship with intentions. Intentions were shown to have moderate to strong significant linear correlations with attitudes (*r*= 0.75, *p*< 0.001) and subjective norms (*r*= 0.75, *p*< 0.001). Necessity, medication to cure/control, and a chronic timeline were all found to have moderate significant negative linear relationships with intention. Attitudes towards discontinuing antidepressants had moderate significant linear correlations with PBC (*r*= 0.59, *p*< 0.001) and necessity (*r* = −0.61, *p*< 0.001).

The results from each step in the hierarchical multiple regression are presented in Table 5. The results showed that the three constructs from the TPB accounted for significant variation in intention scores *F*(3, 157)= 118.04, *p*< 0.001, adj. *R*^2^= 0.62. The addition of salient beliefs (Step 2) to the prediction of intention led to a small but significant increase *R*^2^ change of 0.04, *F*(5, 152)= 3.31, *p* <0.01. There was a minimal change in *R*^2^ when adding past history to the model (Step 3), but this change was not significant *F*(3, 149)= 1.8, *p*= 0.14. The addition of symptom severity (Step 4) and duration of antidepressant treatment (Step 5) did not change *R*^2^.

**Table 5.**
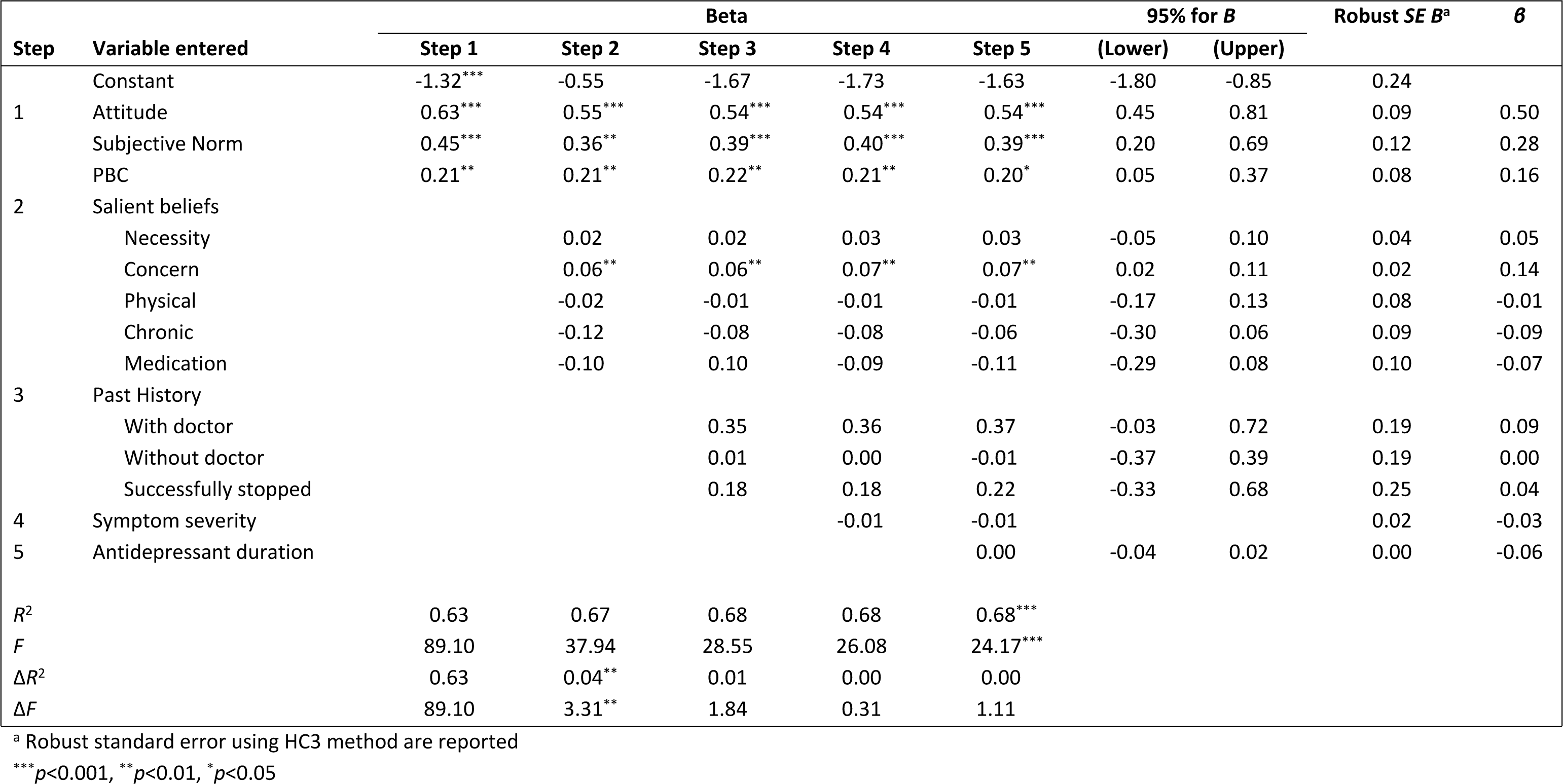
Prediction of intentions using TPB variables, salient beliefs, past history, symptom severity and antidepressant duration.

Each of the models were tested to see whether they were statistically significant in predicting intentions. The full model including all constructs from the TPB, salient beliefs, past history, symptom severity and antidepressant treatment duration to predict intention was statistically significant *R*^2^= 0.69, *F* (13, 147)= 24.17, *p*< 0.001, adjusted *R*^2^= 0.65.

The regression coefficients show that attitude (*B*= 0.54, 95% CI 0.29, 0.78, *p*< 0.001), subjective norm (*B*= 0.39, 95% CI 0.13, 0.66, *p*< 0.001) and PBC (*B*= 0.20, 95% CI 0.03, 0.37, *p*< 0.05) added statistically significantly to predicting intentions. No linear relationships were found between salient beliefs, symptom severity, or current duration of antidepressant treatment. Within the variable of past behaviour, previous attempts to stop taking antidepressants with a doctor’s knowledge and successfully stopping showed a positive linear relationship on intentions to discontinue antidepressants, but were not statistically significant (*B*= 0.37, 95% CI −0.03, 0.72, *p*= 0.06 and *B*= 0.22, 95% CI −0.33 −0.68, *p*= 0.39 respectively). Taking all variables into account, only TPB constructs and concerns maintained their ability to predict intentions towards starting to come off antidepressants.

### Predicting behaviour

The mean intention and PBC scores comparing participants who reduced and did not reduce antidepressant dose are shown in Table 6.

**Table 6.** Comparison of intention and PBC scores between participants who reduced and did not reduce antidepressants at six months.

| Behaviour<br>(N= 151) | Sample<br>N (%) | Intention |  | PBC |  |
| --- | --- | --- | --- | --- | --- |
|  |  | M | SD | M | SD |
| Reduced | 12 (7.95%) | 3.14 | 0.55 | 3.11 | 1.30 |
| Did not reduce | 139 (92.05%) | 2.39 | 1.68 | 3.41 | 1.44 |

The difference in intention scores between those who reduced (mean rank= 101.37) and did not reduce (mean rank = 84.5) was not statistically significant, *U*= 1400.50, *z*= 1.30, *p*= 0.19. There was no statistically significant difference in PBC scores between those who reduced (mean rank = 70.83) and did not reduce (mean rank = 76.45), *U*= 772.00, *z*= −0.42, *p*= 0.67.[53]

The binomial logistic regression model was found to be non-significant, χ^2^(1)= 0.83, *p*= 0.36. Variation in having an appointment with a health professional or not was less than 1%. The model showed no improvement in estimating the probability of having an appointment with a health professional compared to a model that assumed that all cases would be classified as not attending an appointment. The model’s sensitivity was poor in that it did not correctly predict any participants who did have an appointment (n= 60). The specificity of the model was high in that all participants (n= 105) who did not have an appointment with a health professional were correctly predicted not to have had an appointment. The odds of having an appointment increased with stronger intentions towards starting to come off antidepressants, but this finding was not statistically significant, Exp *B*= 1.09, 95% CI 0.91, 1.31, *p*= 0.36.

## Discussion

The aim of this study was to determine whether beliefs and attitudes towards long-term antidepressant use predict intentions to stop treatment; and whether these intentions translate into actual behaviour. As well as conducting an exploratory analysis of survey responses, a further objective was to determine how well participants’ beliefs and attitudes could be explained by an extended model of the TPB.

Overall, most participants had little to no intention to start to come off antidepressants, and fewer than 10% of the sample had started to reduce their antidepressant dose at six months. The full model was found to significantly predict 65% of the variance in intentions towards starting to come off antidepressants; with more favourable attitudes and normative beliefs the strongest predictors on intentions to stop long-term antidepressant treatment.

Most participants believed their depression was chronic, felt taking antidepressants were necessary, and were generally comfortable with taking them. Necessity beliefs about antidepressants appeared to be the most important factor when considering stopping long-term antidepressant use, with over 85% of participants agreeing that taking antidepressants was necessary. Furthermore, the proportion of variance of concerns in predicting attitudes and intentions was small, in line with other research showing that patients may not prioritise concerns about taking antidepressants over the perceived risks of discontinuation.[54−60] Stronger beliefs in the necessity of antidepressants are related to higher levels of adherence in the initial stages of antidepressant treatment; with beliefs in the necessity of antidepressants continuing to increase over time.[22, 61−63] This may explain the findings that stronger beliefs in the chronicity of depression predict fewer intentions to discontinue treatment.

While few participants started to come off antidepressants, an important finding is that subjective norms were significant in predicting intentions to start to come off antidepressants. Most participants stated they would be comfortable if their doctor gave them support and follow up if they were to discontinue treatment, which is supported by existing literature.[57, 59, 64−66] Previous attempts to stop with a doctors’ knowledge showed a positive association towards intentions to stop antidepressants, and 11 out of the 16 participants who successfully started antidepressant discontinuation had a face-to-face appointment with their GP. Ongoing monitoring and review and a positive relationship with the GP is important for patients to receive appropriate guidance and support during the acute and maintenance phase of treatment and could facilitate decision-making around stopping treatment and subsequent discontinuation.[65, 67−69]

As 85% of prescriptions were issued using remote methods, few participants had face-to-face contact with their GP. Requesting prescriptions remotely may limit opportunities for patients to talk about their antidepressant use and potential discontinuation with their GP. Trials have shown that prompting GPs to review their patients’ long-term antidepressant use will result in a proportion of patients discontinuing. In unselected samples, around 6-8% will discontinue antidepressants after practitioner review.[70, 71] If the patients are selected for their willingness to try discontinuing, the rate is higher than 40%, with little risk of relapse of depression, at least up to 12 months after discontinuation.[72]

Patients should be encouraged to attend more face-to-face consultations to discuss management, long-term risks of antidepressant use, and continued support, should they wish to discontinue treatment.[15, 57, 65, 73, 74] Internet and telephone support for discontinuing antidepressants, after primary care practitioner review and advice about tapering the dose, can reduce the risk of depressive and withdrawal symptoms, and conserve mental wellbeing.[72] However, both patients and GPs demonstrate uncertainty about who is responsible for initiating a consultation to review their antidepressant use.[59, 69, 75-77] Furthermore, there is limited guidance on how to initiate discussions around discontinuation or how to manage patients’ fears and uncertainties, with varying levels in GP confidence when listening to and managing patients’ fears and concerns around discontinuing long-term antidepressant use.[64, 77, 78]

### Strengths & Limitations

To the best of our knowledge, no previous research has explored how beliefs and attitudes may predict intentions towards long-term antidepressant discontinuation or examined the strength of the TPB in explaining behaviours regarding antidepressant use. The findings suggest that the utility of the TPB in predicting intentions towards discontinuing long-term antidepressant use is similar to its utility when applied to other health-related behaviours, where it has been shown to explain between 40-49% of the variance in intentions.[79] However, there are some limitations of the TPB that should be considered. While it is acceptable to add additional predictors to the model, they should only be added if they can show a significant proportion of variance in intentions or behaviour in addition to the original constructs of the TPB.[25] Past behaviour is considered one of the strongest predictors of future behaviour, but only if it is performed frequently.[31, 36] In the current study, past history accounted for very little change in the variance of the model in predicting intentions, but could be explained in that antidepressant discontinuation is not frequently performed.[57, 58, 60] The intention-behaviour gap should also be considered.[80] Some participants with little intention to start to come off antidepressants did eventually reduce or stop their antidepressant dose. As the majority of participants that stopped or reduced their dose had an appointment with their GP or a pharmacist, this suggests that a review consultation could act as an implementation intention and may ‘bridge the gap’ between behaviour and intentions.[80–84]

A key limitation of the research was the level of missing data and the use of self-report data to measure current antidepressant duration. Participants self-reported a median continuous antidepressant treatment duration of 11 years, which is considerably higher than the average reported length of treatment in previous research.[71, 85, 86] As some participants stated they did not know how long they had been on antidepressants, this self-reported higher duration of treatment could be based on participants’ best guess rather than prescribing data reported in published data. This, along with many participants responding that they did not know how long they had been taking antidepressants for, is an interesting finding. Despite testing the face validity of the questionnaire through cognitive interviews, many responses for the current study were left blank or were difficult to interpret.

Furthermore, as most participants indicated little to no intention to start to come off antidepressants in the next six months, and less than 10% of participants reduced their antidepressant dose at six months, it was difficult to create a reliable predictive model. Due to missing data, the error may be over-estimated and the power of the model is low, reducing the likelihood that a statistically significant result shows a true effect, and making it difficult to rule out a Type II error.[87] This may be the case for concerns about antidepressants and previous attempts to stop with the doctor’s knowledge, as the results found for these possible predictors were in the direction of a positive association and approached the 5% level of statistical significance (*p*= 0.06 in both cases). Therefore, while the models predicting intentions were more robust than the model predicting behaviour, it is not possible to make reliable inferences about how well the TPB can explain intentions to stop long-term antidepressant treatment.

The sociodemographic characteristics of participants who took part in the study should be considered. Nearly all participants were from a White ethnic group, so the findings may not represent the beliefs and attitudes of patients from ethnic minority backgrounds.

Research[88] suggests that people from ethnic minority backgrounds have weaker beliefs in the biological causes of depression compared to people from a White ethnic background, and have stronger beliefs in the psychosocial causes of depression. This may explain why people from ethnic minority backgrounds are less likely to believe that antidepressants are effective in managing depression,[89] and hold stronger beliefs that antidepressants are addictive.[88] Difficulties in recruiting underrepresented groups to mental health research are unfortunately not uncommon.[90] Overall health-related deprivation patterns are evident in England, with significant health inequalities between the North and the South of the country, which can be explained by socioeconomic deprivation.[91] Participants were recruited through GP practices based in the South and South-West of England. Therefore, the participating practices may not represent those from areas with higher levels of socioeconomic deprivation. Future research needs to explore the beliefs, attitudes, and behavioural intentions towards long-term antidepressant use both between and within different sociodemographic groups.

### Implications for primary care

The NICE guidelines emphasise the need for continued monitoring and review of people on long-term antidepressant treatment.[37] However, few review consultations happen with people who use antidepressants long-term, with the percentage of people reviewed during each year of treatment decreasing over ten years.[75, 92] Reviewing long-term antidepressant use can reduce drug burden, with a primary care pharmacist-led study showing that around 15% of people who had an active review had their antidepressant therapy altered, which led to a reduction in antidepressant prescribing.[71] This emphasises the importance of GPs to invite people who have been on antidepressants for more than two years to a review.[93] However, minimising inappropriate long-term antidepressant use can be challenging for GPs,[77, 78, 94, 95] due to perceived patient demand for antidepressants treatment and a lack of opportunity to have review consultations.[96] Furthermore, people may prefer requesting repeat prescriptions remotely or are ambivalent about the need for a consultation if the GP continues to approve prescriptions without a review.[57, 73] The research suggests a need for a more patient-centred approach to the management of depression in primary care, where the beliefs about depression and treatment preferences are key considerations when formulating a treatment plan.[88, 97, 98] Given strong negative views towards intentions to stop taking antidepressants and concerns around symptoms of withdrawal and relapse during discontinuation,[64, 69, 74, 99-105] further discussions between the patient and GP around beliefs and attitudes towards long-term antidepressants are needed from the outset, so patients can actively consider their intentions towards discontinuing long-term use. Patients need to be aware of the importance of ongoing monitoring and review, so that these conversations with the GP can take place. In turn, regular monitoring and review will help maintain a strong GP-patient relationship, which could facilitate conversations around intentions to start to come off antidepressants. This could give patients greater confidence to start the process of antidepressant discontinuation.

To further facilitate the process of discontinuation, GPs need appropriate guidance and support to help inform patients about the role of antidepressants in managing depression, and how to broach the conversation regarding discontinuation. In addition to informing patients at the start of antidepressant treatment that it should not be considered for life and will need to be managed slowly,[106] further guidance is needed for GPs to help manage patients’ fears and uncertainties about symptoms of withdrawal and relapse and appropriate guidance on the tapering process and successful antidepressant discontinuation. Internet and telephone support combined with practitioner review and advice on tapering off treatment can protect patients against depressive and withdrawal symptoms.[72]

Considering more salient beliefs and attitudes patients may have towards the necessity of long-term antidepressants use means GPs may be able to support patients in formulating a plan for reducing their antidepressant dose that addresses their particular beliefs and mitigates any fears and uncertainties they may have.

Moreover, further research could explore patients’ views about discussing antidepressant discontinuation from other health professionals, such as pharmacists or nurse prescribers.

## Data Availability

The Ethical Approvals provided by the Sponsor (University of Southampton), NHS Research Ethics Committee, and Health Research Authority, states that “Access to raw data and right to publish freely by all investigators in study or by Independent Steering Committee on behalf of all investigators” will not form part of the dissemination plan. Furthermore, participants have not provided consent for the data to be shared anonymously for other ethically approved research in the future.

## Supporting information

S1_Appendix. Study Protocol

S2_Appendix. Questionnaire Survey booklet S3_Appendix. Medical Notes data capture form

S1_Table. Mean scores for beliefs about depression and antidepressant discontinuation S2_Table. Means, standard deviations, and intercorrelations for salient beliefs on attitude S3_Table. Prediction of attitudes towards discontinuation using salient beliefs

S4_Table. Means, standard deviations and intercorrelations for beliefs and attitudes on intentions

## Notes

### Competing Interest Statement

I have read the journal's policy and the authors of this manuscript have the following competing interests: TK, AWG and BS have received grant funding from the NIHR Research for Patient Benefit programme to conduct research into helping people come off inappropriate long-term antidepressants.

### Funding Statement

Yes

### Author Declarations

The study was ethically approved through proportionate review by Yorkshire & The Humber – Leeds East Research Ethics Committee (REC ID: 17/YH/0223). Completion of the questionnaire indicated implied consent. Participants were required to provide written consent for their GP practice to provide medical notes data.

## References

1. Lewis G, Marston L, Duffy L, Freemantle N, Gilbody S, Hunter R, et al. Maintenance or Discontinuation of Antidepressants in Primary Care. N Engl J Med. 2021;385(14):1257–67. doi: 10.1056/nejmoa2106356.

2. Mars B, Heron J, Kessler D, Davies NM, Martin RM, Thomas KH, Gunnell D. Influences on antidepressant prescribing trends in the UK: 1995–2011. Soc Psychiatry Psychiatr Epidemiol. 2017;52(2):193–200. doi: 10.1007/s00127-016-1306-4.

3. Public Health England. Public Health Profiles. Depression: Recorded prevalence (aged 18+) 2019/20 2021 [cited 2021 5th May 2021]. Available from: https://fingertips.phe.org.uk/search/depression#page/3/gid/1/pat/6/par/E12000001/ati/102/are/E06000047/iid/848/age/168/sex/4/cid/4/tbm/1/page-options/ovw-do-0.

4. Kendrick T, Stuart B, Newell C, Geraghty AW, Moore M. Changes in rates of recorded depression in English primary care 2003–2013: Time trend analyses of effects of the economic recession, and the GP contract quality outcomes framework (QOF). J Affect Disord. 2015;180:68–78. Epub 2015/04/17. doi: 10.1016/j.jad.2015.03.040. PubMed PMID: 25881283.

5. NHS Digital. Prescription cost analysis - England: 2018: NHS Digital; 2018 [5th May 2021]. Available from: https://digital.nhs.uk/data-and-information/publications/statistical/prescription-cost-analysis/2018.

6. Ilyas S, Moncrieff J. Trends in prescriptions and costs of drugs for mental disorders in England, 1998-2010. Br J Psychiatry. 2012;200(5):393–8. Epub 2012/03/24. doi: 10.1192/bjp.bp.111.104257. PubMed PMID: 22442100.

7. Moore M, Yuen HM, Dunn N, Mullee MA, Maskell J, Kendrick T. Explaining the rise in antidepressant prescribing: a descriptive study using the general practice research database. BMJ. 2009;339:b3999. Epub 2009/10/17. doi: 10.1136/bmj.b3999. PubMed PMID: 19833707; PubMed Central PMCID: PMCPMC2762496.

8. Verhaak PFM, de Beurs D, Spreeuwenberg P. What proportion of initially prescribed antidepressants is still being prescribed chronically after 5 years in general practice? A longitudinal cohort analysis. BMJ Open. 2019;9(2):e024051. Epub 2019/03/01. doi: 10.1136/bmjopen-2018-024051. PubMed PMID: 30813115; PubMed Central PMCID: PMCPMC6377556.

9. Huijbregts KM, Hoogendoorn A, Slottje P, van Balkom AJ, Batelaan NM. Long-term and short-term antidepressant use in general practice: data from a large cohort in the Netherlands. Psychother Psychosom. 2017;86(6):362–9. doi: 10.1159/000480456.

10. Wilkinson P, Izmeth Z. Continuation and maintenance treatments for depression in older people. Cochrane Database Syst Rev. 2016;9. Epub 2016/09/10. doi: 10.1002/14651858.CD006727.pub2. PubMed PMID: 27609183; PubMed Central PMCID: PMCPMC6457610.

11. McCrea RL, Sammon CJ, Nazareth I, Petersen I. Initiation and duration of selective serotonin reuptake inhibitor prescribing over time: UK cohort study. Br J Psychiatry. 2016;209(5):421–6. Epub 2016/11/03. doi: 10.1192/bjp.bp.115.166975. PubMed PMID: 27539294.

12. Cruickshank G, MacGillivray S, Bruce D, Mather A, Matthews K, Williams B. Cross-sectional survey of patients in receipt of long-term repeat prescriptions for antidepressant drugs in primary care. Mental Health in Family Medicine. 2008;5(2):105–9. Epub 2008/06/01. PubMed PMID: PMC2777559; PubMed Central PMCID: PMCPMC2777559.

13. Hughes S, Cohen D. A systematic review of long-term studies of drug treated and non-drug treated depression. J Affect Disord. 2009;118(1-3):9–18. Epub 2009/03/03. doi: 10.1016/j.jad.2009.01.027. PubMed PMID: 19249104.

14. Zajecka JM. Clinical issues in long-term treatment with antidepressants. J Clin Psychiatry. 2000;61 Suppl 2:20–5. Epub 2000/03/14. PubMed PMID: 10714620.

15. Cartwright C, Gibson K, Read J, Cowan O, Dehar T. Long-term antidepressant use: patient perspectives of benefits and adverse effects. Patient Preference and Adherence. 2016;10:1401–7. Epub 2016/08/17. doi: 10.2147/PPA.S110632. PubMed PMID: 27528803; PubMed Central PMCID: PMCPMC4970636.

16. Ferguson JM. SSRI Antidepressant Medications: Adverse Effects and Tolerability. Primary Care Companion J Clin Psychiatry. 2001;3(1):22–7. Epub 2004/03/12. doi: 10.4088/pcc.v03n0105. PubMed PMID: 15014625; PubMed Central PMCID: PMCPMC181155.

17. Hollon SD, DeRubeis RJ, Andrews PW, Thomson JA. Cognitive Therapy in the Treatment and Prevention of Depression: A Fifty-Year Retrospective with an Evolutionary Coda. Cognit Ther Res. 2021;45(3):402–17. doi: 10.1007/s10608-020-10132-1. PubMed PMID: WOS:000548799700001.

18. Coupland C, Dhiman P, Morriss R, Arthur A, Barton G, Hippisley-Cox J. Antidepressant use and risk of adverse outcomes in older people: population based cohort study. BMJ. 2011;343:d4551. Epub 2011/08/04. doi: 10.1136/bmj.d4551. PubMed PMID: 21810886; PubMed Central PMCID: PMCPMC3149102.

19. Lynch J, Moore M, Moss-Morris R, Kendrick T. Do patients’ illness beliefs predict depression measures at six months in primary care; a longitudinal study. J Affect Disord. 2015;174:665–71. doi: 10.1016/j.jad.2014.12.005. PubMed PMID: WOS:000350003800091.

20. Lynch J, Moore M, Moss-Morris R, Kendrick T. Are patient beliefs important in determining adherence to treatment and outcome for depression? Development of the beliefs about depression questionnaire. J Affect Disord. 2011;133:29–41. Epub 2011/04/22. doi: 10.1016/j.jad.2011.03.019. PubMed PMID: 21507489.

21. Brown C, Dunbar-Jacob J, Palenchar DR, Kelleher KJ, Bruehlman RD, Sereika S, Thase ME. Primary care patients’ personal illness models for depression: a preliminary investigation. Fam Pract. 2001;18(3):314–20. Epub 2001/05/18. doi: 10.1093/fampra/18.3.314. PubMed PMID: 11356741.

22. Aikens JE, Nease DE, Jr., Klinkman MS. Explaining patients’ beliefs about the necessity and harmfulness of antidepressants. Ann Fam Med. 2008;6(1):23–9. Epub 2008/01/16. doi: 10.1370/afm.759. PubMed PMID: 18195311; PubMed Central PMCID: PMCPMC2203394.

23. Read J, Cartwright C, Gibson K, Shiels C, Magliano L. Beliefs of people taking antidepressants about the causes of their own depression. J Affect Disord. 2015;174:150–6. Epub 2014/12/17. doi: 10.1016/j.jad.2014.11.009. PubMed PMID: 25497472.

24. Biesheuvel-Leliefeld KE, Kok GD, Bockting CL, Cuijpers P, Hollon SD, van Marwijk HW, Smit F. Effectiveness of psychological interventions in preventing recurrence of depressive disorder: meta-analysis and meta-regression. J Affect Disord. 2015;174:400–10. Epub 2015/01/02. doi: 10.1016/j.jad.2014.12.016. PubMed PMID: 25553400.

25. Ajzen I. The theory of planned behavior. Organ Behav Hum Decis Process. 1991;50(2):179–211. doi: 10.1016/0749-5978(91)90020-T.

26. Creswell JW, Plano Clark VL. Designing and Conducting Mixed Methods Research. 3rd ed. Thousand Oaks, CA: SAGE Publications; 2017.

27. Bishop FL. Using mixed methods research designs in health psychology: An illustrated discussion from a pragmatist perspective. Br J Health Psychol. 2015;20(1):5–20. Epub 2014/11/19. doi: 10.1111/bjhp.12122. PubMed PMID: 25405549.

28. British Medical Association, Royal Pharmaceutical Society of Great Britain. British National Formulary: 58. London: Pharmaceutical Press; 2009.

29. O’Neil M, Payne C, Read J. Read Codes Version 3: a user led terminology. Methods Inf Med. 1995;34(1-2):187–92. Epub 1995/03/01. PubMed PMID: 9082130.

30. NHS Health Research Authority. Applying a proportionate approach to the process of seeking consent: HRA Guidance. In: Health Research Authority, editor. United Kingdom2018.

31. Conner M, Armitage CJ. Extending the theory of planned behavior: A review and avenues for further research. J Appl Soc Psychol. 1998;28(15):1429–64. doi: 10.1111/j.1559-1816.1998.tb01685.x. PubMed PMID: WOS:000075600100008.

32. Fishbein M. Attitude and the predictions of behaviour. In: Fishbein M, editor. Readings in attitude theory and measurement. New York, NY: John Wiley & Sons; 1967.

33. Lynch J, Kendrick T, Moore M, Johnston O, Smith PWF. Patients’ beliefs about depression and how they relate to duration of antidepressant treatment: use of a US measure in a UK primary care population. Primary Care Mental Health. 2006;4(3):207–17. PubMed PMID: 105927789. Language: English. Entry Date: 20080118. Revision Date: 20150820. Publication Type: Journal Article.

34. Reeve E, To J, Hendrix I, Shakib S, Roberts MS, Wiese MD. Patient barriers to and enablers of deprescribing: a systematic review. Drugs Aging. 2013;30(10):793–807. Epub 2013/08/06. doi: 10.1007/s40266-013-0106-8. PubMed PMID: 23912674.

35. Horne R, Weinman J. Patients’ beliefs about prescribed medicines and their role in adherence to treatment in chronic physical illness. J Psychosom Res. 1999;47(6):555–67. Epub 2000/02/08. doi: 10.1016/s0022-3999(99)00057-4. PubMed PMID: 10661603.

36. Sutton S. The past predicts the future: Interpreting behaviour–behaviour relationships in social psychological models of health behaviour. In: Rutter DR, Quine L, editors. Social psychology and health: European perspectives. Brookfield, VT: Avebury/Ashgate Publishing Co; 1994. p. 71-88.

37. National Institute for Health and Care Excellence. Depression in adults: recognition and management 2009 [cited 2017 15th November 2017]. Available from: www.nice.org.uk/guidance/cg90.

38. Hagger MS. Retired or not, the theory of planned behaviour will always be with us. Health Psychol Rev. 2015;9(2):125–30. Epub 2015/05/13. doi: 10.1080/17437199.2015.1034470. PubMed PMID: 25966053.

39. Sniehotta FF, Presseau J, Araújo-Soares V. Time to retire the theory of planned behaviour. Health Psychol Rev. 2014;8(1):1–7. Epub 2014/07/24. doi: 10.1080/17437199.2013.869710. PubMed PMID: 25053004.

40. Sutton S. Determinants of health-related behaviours: Theoretical and methodological issues. In: Sutton S, Baum A, Johnston M, editors. The Sage Handbook of Health Psychology. London: SAGE Publications; 2004. p. 94-126.

41. Francis JJ, Eccles MP, Johnston M, Walker A, Grimshaw J, Foy R, et al. Constructing questionnaires based on the theory of planned behaviour. A manual for health services researchers. 2004;2010:2–12.

42. Dewar-Haggart R. Exploring beliefs, attitudes, and behavioural intentions towards long-term antidepressant use in the management of people with depression in primary care: a mixed methods study [Doctoral Thesis]: University of Southampton; 2022.

43. Ajzen I. Constructing a Theory of Planned Behaviour Questionnaire2006.

44. Reeve E, Shakib S, Hendrix I, Roberts MS, Wiese MD. Development and validation of the patients’ attitudes towards deprescribing (PATD) questionnaire. Int J Clin Pharm. 2013;35(1):51–6. Epub 2012/10/12. doi: 10.1007/s11096-012-9704-5. PubMed PMID: 23054137.

45. Horne R, Weinman J, Hankins M. The beliefs about medicines questionnaire: The development and evaluation of a new method for assessing the cognitive representation of medication. Psychol Health. 1999;14(1):1–24. doi: 10.1080/08870449908407311. PubMed PMID: WOS:000079471100001.

46. Kroenke K, Strine TW, Spitzer RL, Williams JB, Berry JT, Mokdad AH. The PHQ-8 as a measure of current depression in the general population. J Affect Disord. 2009;114(1-3):163–73. Epub 2008/08/30. doi: 10.1016/j.jad.2008.06.026. PubMed PMID: 18752852.

47. Kroenke K, Spitzer RL, Williams JB. The PHQ-9: validity of a brief depression severity measure. J Gen Intern Med. 2001;16(9):606–13. Epub 2001/09/15. doi: 10.1046/j.1525-1497.2001.016009606.x. PubMed PMID: 11556941; PubMed Central PMCID: PMCPMC1495268.

48. Kroenke K, Spitzer RL. The PHQ-9: A new depression diagnostic and severity measure. Psychiatric Annals. 2002;32(9):509–15. doi: 10.3928/0048-5713-20020901-06. PubMed PMID: WOS:000178070800004.

49. Cohen J. Statistical Power Analysis for the Behavioral Sciences. 2nd ed. New York, NY: Lawrence Erlbaum Associates; 1988.

50. Green SB. How Many Subjects Does It Take to Do a Regression-Analysis. Multivariate Behavioral Research. 1991;26(3):499–510. doi: 10.1207/s15327906mbr2603_7. PubMed PMID: WOS:A1991GT15700007.

51. Tabachnick BG, Fidell LS. Using Multivariate Statistics: Pearson New International Edition. Harlow: Pearson Education UK; 2013.

52. IBM Corp. IBM SPSS Statistics for Windows, Version 26.0. Armonk, NY: IBM Corp; 2019.

53. Mann HB, Whitney DR. On a test of whether one of two random variables is stochastically larger than the other. The Annals of Mathematical Statistics. 1947:50–60.

54. Eveleigh R, Speckens A, van Weel C, Oude Voshaar R, Lucassen P. Patients’ attitudes to discontinuing not-indicated long-term antidepressant use: barriers and facilitators. Therapeutic Advances in Psychopharmacology. 2019;9:1–9. Epub 2019/09/14. doi: 10.1177/2045125319872344. PubMed PMID: 31516691; PubMed Central PMCID: PMCPMC6724488.

55. Malpass A, Shaw A, Sharp D, Walter F, Feder G, Ridd M, Kessler D. “Medication career” or “moral career”? The two sides of managing antidepressants: a meta-ethnography of patients’ experience of antidepressants. Soc Sci Med. 2009;68(1):154–68. Epub 2008/11/18. doi: 10.1016/j.socscimed.2008.09.068. PubMed PMID: 19013702.

56. Wouters H, Van Dijk L, Van Geffen EC, Gardarsdottir H, Stiggelbout AM, Bouvy ML. Primary-care patients’ trade-off preferences with regard to antidepressants. Psychol Med. 2014;44(11):2301–8. Epub 2014/01/09. doi: 10.1017/S0033291713003103. PubMed PMID: 24398071.

57. Leydon GM, Rodgers L, Kendrick T. A qualitative study of patient views on discontinuing long-term selective serotonin reuptake inhibitors. Fam Pract. 2007;24(6):570–5. Epub 2007/11/23. doi: 10.1093/fampra/cmm069. PubMed PMID: 18032401.

58. Verbeek-Heida PM, Mathot EF. Better safe than sorry -why patients prefer to stop using selective serotonin reuptake inhibitor (SSRI) antidepressants but are afraid to do so: results of a qualitative study. Chronic Illness. 2006;2(2):133–42. doi: 10.1179/174592006x111003.

59. Bosman RC, Huijbregts KM, Verhaak PF, Ruhe HG, van Marwijk HW, van Balkom AJ, Batelaan NM. Long-term antidepressant use: a qualitative study on perspectives of patients and GPs in primary care. Br J Gen Pract. 2016;66(651):e708–19. Epub 2016/08/17. doi: 10.3399/bjgp16X686641. PubMed PMID: 27528709; PubMed Central PMCID: PMCPMC5033307.

60. Dickinson R, Knapp P, House AO, Dimri V, Zermansky A, Petty D, et al. Long-term prescribing of antidepressants in the older population: a qualitative study. Br J Gen Pract. 2010;60(573):e144–55. Epub 2010/04/01. doi: 10.3399/bjgp10X483913. PubMed PMID: 20353660; PubMed Central PMCID: PMCPMC2845505.

61. Aikens JE, Klinkman MS. Changes in patients’ beliefs about their antidepressant during the acute phase of depression treatment. Gen Hosp Psychiatry. 2012;34(3):221–6. Epub 2012/02/14. doi: 10.1016/j.genhosppsych.2012.01.004. PubMed PMID: 22325627.

62. Aikens JE, Nease Jr DE, Nau DP, Klinkman MS, Schwenk TL. Adherence to maintenance-phase antidepressant medication as a function of patient beliefs about medication. Ann Fam Med. 2005;3(1):23–30. Epub 2005/01/27. doi: 10.1370/afm.238. PubMed PMID: 15671187; PubMed Central PMCID: PMCPMC1466796.

63. Brown C, Battista DR, Bruehlman R, Sereika SS, Thase ME, Dunbar-Jacob J. Beliefs about antidepressant medications in primary care patients: Relationship to self-reported adherence. Med Care. 2005;43(12):1203–7. doi: 10.1097/01.mlr.0000185733.30697.f6. PubMed PMID: WOS:000233711100007.

64. Wentink C, Huijbers MJ, Lucassen PL, van der Gouw A, Kramers C, Spijker J, Speckens AE. Enhancing shared decision making about discontinuation of antidepressant medication: a concept-mapping study in primary and secondary mental health care. Br J Gen Pract. 2019;69(688):e777–85. Epub 2019/09/25. doi: 10.3399/bjgp19X706001. PubMed PMID: 31548298; PubMed Central PMCID: PMCPMC6758920.

65. Maund E, Dewar-Haggart R, Williams S, Bowers H, Geraghty AWA, Leydon G, et al. Barriers and facilitators to discontinuing antidepressant use: A systematic review and thematic synthesis. J Affect Disord. 2019;245:38–62. Epub 2018/10/27. doi: 10.1016/j.jad.2018.10.107. PubMed PMID: 30366236.

66. Backenstrass M, Joest K, Rosemann T, Szecsenyi J. The care of patients with subthreshold depression in primary care: Is it all that bad? A qualitative study on the views of general practitioners and patients. BMC Health Serv Res. 2007;7(190). doi: 10.1186/1472-6963-7-190.

67. Gibson K, Cartwright C, Read J. Patient-centered perspectives on antidepressant use: a narrative review. Int J Ment Health. 2014;43(1):81–99. doi: 10.2753/IMH0020-7411430105.

68. Badger F, Nolan P. Concordance with antidepressant medication in primary care. Nurs Stand. 2006;20(52):35–40. Epub 2006/09/23. doi: 10.7748/ns2006.09.20.52.35.c4492. PubMed PMID: 16989339.

69. Scholten W, Batelaan N, Van Balkom A. Barriers to discontinuing antidepressants in patients with depressive and anxiety disorders: a review of the literature and clinical recommendations. Therapeutic Advances in Psychopharmacology. 2020;10:1–10. Epub 2020/06/25. doi: 10.1177/2045125320933404. PubMed PMID: 32577215; PubMed Central PMCID: PMCPMC7290254.

70. Eveleigh R, Grutters J, Muskens E, Oude Voshaar R, van Weel C, Speckens A, Lucassen P. Cost-utility analysis of a treatment advice to discontinue inappropriate long-term antidepressant use in primary care. Fam Pract. 2014;31(5):578–84. Epub 2014/08/15. doi: 10.1093/fampra/cmu043. PubMed PMID: 25121977.

71. Johnson CF, Macdonald HJ, Atkinson P, Buchanan AI, Downes N, Dougall N. Reviewing long-term antidepressants can reduce drug burden: a prospective observational cohort study. Br J Gen Pract. 2012;62(604):e773–9. Epub 2012/12/06. doi: 10.3399/bjgp12X658304. PubMed PMID: 23211181; PubMed Central PMCID: PMCPmc3481518.

72. Kendrick T, Stuart B, Bowers H, Sadeghi M, Page H, Dowrick C, et al. Internet and telephone support for discontinuing long-term antidepressants: cluster randomized trial. REDUCE open pragmatic effectivenes trial in UK primary care. In Press.

73. Gask L, Rogers A, Oliver D, May C, Roland M. Qualitative study of patients’ perceptions of the quality of care for depression in general practice. Br J Gen Pract. 2003;53(489):278–83. Epub 2003/07/26. PubMed PMID: 12879827; PubMed Central PMCID: PMCPMC1314569.

74. Wentink C, Huijbers MJ, Lucassen P, Kramers C, Akkermans R, Adang E, et al. Discontinuation of antidepressant medication in primary care supported by monitoring plus mindfulness-based cognitive therapy versus monitoring alone: design and protocol of a cluster randomized controlled trial. BMC Fam Pract. 2019;20(105):1–9. Epub 2019/07/28. doi: 10.1186/s12875-019-0989-5. PubMed PMID: 31349796; PubMed Central PMCID: PMCPMC6660713.

75. Sinclair JE, Aucott LS, Lawton K, Reid IC, Cameron IM. The monitoring of longer term prescriptions of antidepressants: observational study in a primary care setting. Fam Pract. 2014;31(4):419–26. Epub 2014/05/23. doi: 10.1093/fampra/cmu019. PubMed PMID: 24850795.

76. Middleton N, Gunnell D, Whitley E, Dorling D, Frankel S. Secular trends in antidepressant prescribing in the UK, 1975-1998. J Public Health Med. 2001;23(4):262–7. Epub 2002/03/05. doi: 10.1093/pubmed/23.4.262. PubMed PMID: 11873886.

77. Bowers HM, Williams SJ, Geraghty AWA, Maund E, O’Brien W, Leydon G, et al. Helping people discontinue long-term antidepressants: views of health professionals in UK primary care. BMJ Open. 2019;9(7):e027837. Epub 2019/07/07. doi: 10.1136/bmjopen-2018-027837. PubMed PMID: 31278099; PubMed Central PMCID: PMCPMC6615882.

78. Kelly D, Graffi J, Noonan M, Green P, McFarland J, Hayes P, Glynn L. Exploration of GP perspectives on deprescribing antidepressants: a qualitative study. BMJ Open. 2021;11(4):e046054. Epub 2021/04/07. doi: 10.1136/bmjopen-2020-046054. PubMed PMID: 33820792; PubMed Central PMCID: PMCPMC8030471.

79. McEachan RRC, Conner M, Taylor NJ, Lawton RJ. Prospective prediction of health-related behaviours with the Theory of Planned Behaviour: a meta-analysis. Health Psychol Rev. 2011;5(2):97–144. doi: 10.1080/17437199.2010.521684. PubMed PMID: WOS:000297300600001.

80. Sheeran P. Intention-Behavior Relations: A Conceptual and Empirical Review. European Review of Social Psychology. 2002;12(1):1–36. doi: 10.1080/14792772143000003.

81. Sutton S. Predicting and explaining intentions and behavior: How well are we doing? J Appl Soc Psychol. 1998;28(15):1317–38.

82. Sniehotta FF, Scholz U, Schwarzer R. Bridging the intention–behaviour gap: Planning, self-efficacy, and action control in the adoption and maintenance of physical exercise. Psychol Health. 2005;20(2):143–60. doi: 10.1080/08870440512331317670.

83. Gollwitzer PM. Goal achievement: The role of intentions. European Review of Social Psychology. 1993;4(1):141–85. doi: 10.1080/14792779343000059.

84. Rise J, Thompson M, Verplanken B. Measuring implementation intentions in the context of the theory of planned behavior. Scand J Psychol. 2003;44(2):87–95. doi: 10.1111/1467-9450.00325.

85. Petty DR, House A, Knapp P, Raynor T, Zermansky A. Prevalence, duration and indications for prescribing of antidepressants in primary care. Age Ageing. 2006;35(5):523–6. Epub 2006/05/13. doi: 10.1093/ageing/afl023. PubMed PMID: 16690637.

86. Sehmi R, Smith N, Nguyen A, McManus S. Trends in long-term prescribing of dependence forming medicines. London: PHRC/NatCen; 2019.

87. Eekhout I, de Vet HCW, Twisk JWR, Brand JPL, de Boer MR, Heymans MW. Missing data in a multi-item instrument were best handled by multiple imputation at the item score level. J Clin Epidemiol. 2014;67(3):335–42. doi: 10.1016/j.jclinepi.2013.09.009. PubMed PMID: WOS:000330752400013.

88. Givens JL, Houston TK, Van Voorhees BW, Ford DE, Cooper LA. Ethnicity and preferences for depression treatment. Gen Hosp Psychiatry. 2007;29(3):182–91. doi: 10.1016/j.genhosppsych.2006.11.002.

89. Cooper LA, Gonzales JJ, Gallo JJ, Rost KM, Meredith LS, Rubenstein LV, et al. The acceptability of treatment for depression among African-American, Hispanic, and white primary care patients. Med Care. 2003;41(4):479–89. doi: 10.1097/01.MLR.0000053228.58042.E4.

90. Woodall A, Morgan C, Sloan C, Howard L. Barriers to participation in mental health research: are there specific gender, ethnicity and age related barriers? BMC Psychiatry. 2010;10(1):1–10.

91. Kontopantelis E, Mamas MA, van Marwijk H, Ryan AM, Buchan IE, Ashcroft DM, Doran T. Geographical epidemiology of health and overall deprivation in England, its changes and persistence from 2004 to 2015: a longitudinal spatial population study. J Epidemiol Community Health. 2018;72(2):140–7. doi: 10.1136/jech-2017-209999. PubMed PMID: WOS:000423984300008.

92. Middleton DJ, Cameron IM, Reid IC. Continuity and monitoring of antidepressant therapy in a primary care setting. Qual Prim Care. 2011;19(2):109–13. Epub 2011/05/18. PubMed PMID: 21575333.

93. Kendrick T. Long-term antidepressant treatment: time for a review? Prescriber. 2015;26(19):7–10. doi: 10.1002/psb.1389.

94. Eveleigh R, Speckens A, van Weel C, Oude Voshaar R, Lucassen P. Patients’ attitudes to discontinuing inappropriate long-term antidepressant use: barriers and facilitators. In: Eveleigh R, editor. Inappropriate long-term antidepressant use in primary care: a challenge to change. Netherlands: Roadboud Universiteit Nijmegen; 2015. p. 75–90.

95. Donald M, Partanen R, Sharman L, Lynch J, Dingle GA, Haslam C, van Driel M. Long-term antidepressant use in general practice: a qualitative study of GPs’ views on discontinuation. Br J Gen Pract. 2021;71(708):e508–e16. Epub 2021/04/21. doi: 10.3399/BJGP.2020.0913. PubMed PMID: 33875415; PubMed Central PMCID: PMCPMC8074642.

96. Johnson CF, Williams B, MacGillivray SA, Dougall NJ, Maxwell M. ’Doing the right thing’: factors influencing GP prescribing of antidepressants and prescribed doses. BMC Fam Pract. 2017;18(72). Epub 2017/06/19. doi: 10.1186/s12875-017-0643-z. PubMed PMID: 28623894; PubMed Central PMCID: PMCPMC5473964.

97. Karasz A, Dowrick C, Byng R, Buszewicz M, Ferri L, Hartman TCO, et al. What we talk about when we talk about depression: doctor-patient conversations and treatment decision outcomes. Br J Gen Pract. 2012;62(594):e55–e63. doi: 10.3399/bjgp12X616373.

98. Gum AM, Areán PA, Hunkeler E, Tang L, Katon W, Hitchcock P, et al. Depression treatment preferences in older primary care patients. The Gerontologist. 2006;46(1):14–22. doi: 10.1093/geront/46.1.14.

99. Van Leeuwen E, van Driel ML, Horowitz MA, Kendrick T, Donald M, De Sutter AI, et al. Approaches for discontinuation versus continuation of long-term antidepressant use for depressive and anxiety disorders in adults. Cochrane Database Syst Rev. 2021;4. Epub 2021/04/23. doi: 10.1002/14651858.CD013495.pub2. PubMed PMID: 33886130; PubMed Central PMCID: PMCPMC8092632.

100. Hengartner MP, Schulthess L, Sorensen A, Framer A. Protracted withdrawal syndrome after stopping antidepressants: a descriptive quantitative analysis of consumer narratives from a large internet forum. Therapeutic Advances in Psychopharmacology. 2020;10:1–13. Epub 2021/01/26. doi: 10.1177/2045125320980573. PubMed PMID: 33489088; PubMed Central PMCID: PMCPMC7768871.

101. Groot PC, van Os J. Outcome of antidepressant drug discontinuation with tapering strips after 1-5 years. Therapeutic Advances in Psychopharmacology. 2020;10:1–8. Epub 2020/09/22. doi: 10.1177/2045125320954609. PubMed PMID: 32953040; PubMed Central PMCID: PMCPMC7476339.

102. Horowitz MA, Taylor D. Tapering of SSRI treatment to mitigate withdrawal symptoms. The Lancet Psychiatry. 2019;6(6):538–46. doi: 10.1016/s2215-0366(19)30032-x.

103. Henssler J, Heinz A, Brandt L, Bschor T. Antidepressant Withdrawal and Rebound Phenomena. Deutsches Ärzteblatt International. 2019;116(20):355−61. Epub 2019/07/11. doi: 10.3238/arztebl.2019.0355. PubMed PMID: 31288917; PubMed Central PMCID: PMCPMC6637660.

104. Batelaan NM, Bosman RC, Muntingh A, Scholten WD, Huijbregts KM, van Balkom A. Risk of relapse after antidepressant discontinuation in anxiety disorders, obsessive-compulsive disorder, and post-traumatic stress disorder: systematic review and meta-analysis of relapse prevention trials. BMJ. 2017;358:j3927. Epub 2017/09/15. doi: 10.1136/bmj.j3927. PubMed PMID: 28903922.

105. Kendrick T, Geraghty AWA, Bowers H, Stuart B, Leydon G, May C, et al. REDUCE (Reviewing long-term antidepressant use by careful monitoring in everyday practice) internet and telephone support to people coming off long-term antidepressants: protocol for a randomised controlled trial. Trials. 2020;21(419):1–15. doi: 10.1186/s13063-020-04338-7. PubMed PMID: WOS:000536960100002.

106. Kendrick T. Strategies to reduce use of antidepressants. Br J Clin Pharmacol. 2021;87(1):23–33. doi: 10.1111/bcp.14475.

